# Preservation of neutralizing antibody function in COVID-19 convalescent plasma treated using a riboflavin and ultraviolet light-based pathogen reduction technology

**DOI:** 10.1101/2021.02.18.21251437

**Authors:** Susan Yonemura, Lindsay Hartson, Taru Dutt, Marcela Henao-Tamayo, Raymond Goodrich, Susanne Marschner

## Abstract

**Background and Objective:** Convalescent plasma (CP) has been embraced as a safe therapeutic option for coronavirus disease 2019 (COVID-19) while other treatments are developed. However, transfusion-transmitted disease is a risk, particularly in regions with high endemic prevalence of transfusion-transmissible diseases. Pathogen reduction can mitigate this risk; thus, the objective of this study was to evaluate the effect of riboflavin and ultraviolet light (R+UV) pathogen reduction technology on the functional properties of CCP.

**Materials and Methods:** CCP units (n = 6) from recovered COVID-19 research donors were treated with R+UV. Pre- and post-treatment samples were tested for coagulation factor and immunoglobulin retention. Antibody binding to spike protein receptor binding domain (RBD), S1, and S2 epitopes of SARS-CoV-2 was assessed by ELISA.

Neutralizing antibody (nAb) function was assessed by pseudovirus reporter viral particle neutralization (RVPN) assay and plaque reduction neutralization test (PRNT).

**Results:** Mean retention of coagulation factors was ≥ 70% while retention of immunoglobulins was 100%. Starting nAb titers were low, but PRNT_50_ titers did not differ between pre- and post-treatment samples. No statistically significant differences were detected in levels of IgG (*P* ≥ 0.3665) and IgM (*P* ≥ 0.1208) antibodies to RBD, S1, and S2 proteins before and after treatment.

**Conclusion:** R+UV PRT effects on coagulation factors were similar to previous reports, but no significant effects were observed on immunoglobulin concentration and antibody function. SARS-CoV-2 nAb function in COVID-19 convalescent plasma is conserved following R+UV PRT treatment.

## Introduction

The coronavirus disease-2019 (COVID-19) pandemic bears testimony to the risk presented by emerging infectious diseases (EID). Few treatment options are available when novel viruses first arise, but the use of convalescent plasma (CP) may be an expedient therapeutic approach until other medical countermeasures become widely available. CP is a treatment in which putatively antibody-rich plasma is taken from those recovered from the disease and transfused to provide passive immunity to infected patients or susceptible individuals. Case reports of effective use of CP date back to the 1918 influenza pandemic [1] and more recently to EID outbreaks including severe acute respiratory syndrome (SARS) [2, 3], Middle East respiratory syndrome (MERS) [4], H1N1 influenza [5], and Ebola virus disease (EVD) [6]. In the current pandemic COVID-19 CP (CCP) has demonstrated safety with minimal side effects [7], though controlled clinical efficacy data is only beginning to come in [8-10].

While the most effective protocols for treatment with CCP are yet to be defined, plasma transfusion is a routine medical procedure available globally. However, as with any blood product, there is a risk of transmitting bloodborne pathogens with CCP transfusion. The causative agent for COVID-19, severe acute respiratory syndrome coronavirus-2 (SARS-CoV-2), is itself not believed to be transfusion-transmissible [11]. Yet the possibility of co-infections is present, particularly in regions with a high endemic prevalence of other infectious diseases [12]. Pathogen reduction technology (PRT) treatment of CCP is a measure that can be taken to maintain the safety of the blood supply while providing potential benefits to COVID-19 patients.

PRT systems have been developed over the past decades as a proactive means to reduce the residual risk of transfusion-transmitted infections that continues to exist despite the implementation of routine blood safety practices such as donor questionnaires, travel deferrals, and viral screening tests [13, 14]. Donor infections could escape these blood safety measures for a number of reasons, including a “window period” donation where the viral load has not yet reached the detection limit of screening tests, a lack of testing capability for particular infectious agents, or an unfavorable cost-benefit ratio for continuing to implement more and more tests. PRT provides a broad-spectrum means to reduce pathogen loads and inhibit infectivity by disrupting the microorganism’s ability to replicate. Commercial PRT systems use chemicals, ultraviolet (UV) light, or the combination of a photosensitizer and UV light to inactivate pathogens, but pathogen kill must be balanced to preserve the blood product quality [15]. Recently a PRT system based upon riboflavin and UV light (R+UV) has been reported to be effective in inactivating SARS-CoV-2 [16, 17]; the work described herein evaluates the effect of R+UV treatment on functional properties of CCP.

## Methods

### COVID-19 Convalescent Plasma Collection

CCP was provided by an accredited blood center specializing in biomaterial collections for research (Key Biologics, Memphis, TN, USA). CCP was collected by apheresis under an IRB-approved protocol from donors determined to have recovered from COVID-19 and was shipped to Colorado State University. All products were placed into frozen storage at ≤ −20 °C upon receipt until needed for further processing.

### Riboflavin and UV Light Pathogen Reduction Treatment

CCP units were treated using a R+UV PRT system (Mirasol^®^ Pathogen Reduction Technology, Terumo Blood and Cell Technologies, Lakewood, CO, USA) as previously described [18]. Briefly, thawed CCP units were transferred to an illumination bag and mixed with 35 mL of riboflavin solution (500 µmol/L riboflavin in 0.9% sodium chloride, pH 4.0 to 5.0 [Terumo Blood and Cell Technologies, Larne, Ireland]). The prepared units were then placed into the UV illumination device (Terumo Blood and Cell Technologies, Lakewood, CO, USA) and exposed to 6.24 J/mL of energy. Samples for analysis were taken prior to the addition of riboflavin solution (Post-Collect), after addition of riboflavin (Pre-Treat), and after UV illumination (Post-Treat). Sample aliquots were stored frozen (≤ −20°C) in cryovials until testing. CCP units were analyzed for selected coagulation factors, immunoglobulins, and SARS-CoV-2 antibody binding and neutralizing activity.

### Plasma Protein Assays

Coagulation factors were tested at Terumo Blood and Cell Technologies (Lakewood, CO, USA) using the STA Compact Max (Diagnostica Stago US, Parsippany, NJ, USA). Chromogenic assays were used to measure factor VIII activity (Chromogenix Coamatic^®^ Factor VIII reagent, diaPharma, Group, Inc., West Chester, OH, USA) and antithrombin III activity (STA^®^-Stachrom^®^ AT III reagent, Diagnostica Stago). An immuno-turbidimetric method was used to assess von Willebrand factor antigen activity (STA^®^ Liatest^®^ VWF:Ag). Clotting assays included fibrinogen (STA^®^ Fibrinogen 5), Protein C (STA^®^-Staclot^®^ Protein C) and Protein S (STA^®^-Staclot^®^ Protein S). The performance of the STA Compact Max instrument has been qualified for intra-run and total precision for all assays performed.

Plasma immunoglobulins and IgG subclasses were measured by standard quantitative nephelometry (IgG, IgA, IgM at UC Health Anschutz, Aurora, CO, USA; IgG subclasses at ARUP Laboratories, Salt Lake City, UT, USA). The reference laboratories performing immunoglobulin analysis are accredited by the College of American Pathologists (CAP) and maintain Clinical Laboratory Improvement Amendments (CLIA) certification.

### SARS-CoV-2 Functional Assays

An enzyme-linked immunosorbent assay (ELISA) was performed at Colorado State University to test CCP samples and a negative control (normal plasma sample) for antibody binding to the SARS-CoV-2 spike protein receptor-binding domain (RBD) and epitopes associated with the spike protein subunits S1 and S2 (catalog numbers 40592-V08H, 40591-V08H, and 40590-V08B, Sino Biological US Inc., Wayne, PA, USA). The protocol for ELISA was adapted from Robbiani et al. [19] with a few modifications. Briefly, high binding 96-half-well microplates (Corning Life Sciences, Tewksbury, MA, USA) were coated with 50 ng S1, S2, or RBD protein prepared in PBS and incubated overnight at 4 °C. On the next day, the plates were washed 5 times with 180 µL wash solution (PBS + 0.05% Tween 20) and non-specific interactions were blocked using 180 µL blocking buffer (PBS + 0.05% Tween 20 + 2% BSA + 2% normal goat serum [Jackson ImmunoResearch Inc., West Grove, PA, USA]). After 2 hours the plates were washed and different CCP sample dilutions prepared in blocking buffer were added to the wells and incubated for 1 hour. Plates were then washed and incubated for 1 hour with horseradish peroxidase (HRP) conjugated anti-human IgG or anti-human IgM secondary antibodies (Jackson ImmunoResearch Inc.) prepared in blocking buffer (1:10,000 dilution). The colorimetric substrate was developed with the addition of 100 µL TMB substrate (Thermo Fisher Scientific, Rockford, IL, USA), and the reaction was stopped by adding 50 µL 1M sulfuric acid. Absorbance was measured at 450 nm using a BioTek Synergy 2 plate reader (BioTek Instruments Inc., Winooski, VT, USA).

The neutralizing activity of CCP samples was evaluated by two assays, a pseudovirus reporter viral particle neutralization (RVPN) assay and a plaque reduction neutralization test (PRNT). The RVPN assay was performed at Vitalant Research Institute (VRI, San Francisco, CA, USA) as previously described [20, 21]. In brief, a vesicular stomatitis virus (VSV)-firefly luciferase pseudotype modified to express the SARS-CoV-2 spike protein was mixed with four-fold dilutions of heat inactivated CCP. A positive serum control and a negative serum control were also prepared. After incubation for one hour at 37 °C the preparations were used to infect reporter cells that were plated into black 96-well tissue culture treated plates. The reporter cells were lysed after 24 hours at 37 °C and removal of the supernatant. Luciferase activity was measured to determine the RVPN result. NT_50_ titers were estimated by calculating percentages of the no serum control and performing non-linear regression. Titers measuring < 40 are deemed to lack nAbs.

The PRNT assay was conducted in a biosafety level 3 (BSL-3) laboratory at the Colorado State University Infectious Disease Research Center (Fort Collins, CO, USA). CCP samples were heat inactivated for 30 minutes at 56 °C and serial 2-fold dilutions were prepared in a 96-well plate (Greiner Bio One, Monroe, NC, USA). Viral stock (strain hCoV-19/USA/WA1/2020, BEI Resources, Manassas, VA, USA) containing approximately 200 plaque-forming units (pfu) per 0.1 mL was added to each well containing plasma dilutions. Following an incubation period at 37 °C in a 5% CO_2_ incubator, 6-well plates (Greiner Bio One) containing recently confluent Vero cells (ATCC, Manassas, VA, USA) were inoculated with the virus-plasma mixtures. After a second incubation period at 37 °C, 2 mL of overlay (2X MEM with 4% FBS [Peak Serum, Wellington, CO, USA] and agarose) was added to each well. After 24 hours incubation at 37 °C a second overlay containing neutral red (Millipore Sigma, ST. Louis, MO, USA) was dispensed into each well and the number of plaques was counted 48 to 72 hours after initial inoculation. The highest dilution of plasma that inhibited plaque formation by 50% (PRNT_50_) was determined based upon the titer of the viral stock and the number of plaques present at each dilution. Donors with PRNT_50_ titers of less than or equal to 1:20 are considered negative for nAbs.

### Statistical Analysis

Descriptive statistics including the mean and standard deviation were calculated for all continuous parameters. To assess the effect of R+UV PRT treatment, Pre-Treat samples were used as the basis for comparison rather than Post-Collect samples in order to account for dilution with riboflavin solution. Protein retention percentages were calculated by taking the ratio of Post-Treat to Pre-Treat values for each sample pair and multiplying by 100. ELISA results were analyzed by plotting optical density measurements by dilution and calculating the area under the curve (AUC) using the trapezoid method.

Comparisons for parameters passing the Shapiro-Wilk test for normality were performed using a paired, two-tailed t test where statistical significance was defined as α < 0.05. Data sets exhibiting a non-normal distribution were evaluated non-parametrically using a Wilcoxon matched-pairs signed rank test. Statistical analysis was performed using Prism 8 for Windows (GraphPad Software, Inc., San Diego, CA, USA).

## Results

CCP was collected from 6 donors with demographics as described in Table 1. All 6 units met the incoming product specifications for the R+UV PRT process and were successfully treated. Protein retention analysis (Table 2) demonstrated that although there was a statistically significant treatment effect for the coagulation factors, retention was on the order of 70% or better. Of note is that the immunoglobulin concentrations, including those for IgG subclasses, were unaffected by R+UV treatment as demonstrated by retention remaining at 100%.

**Table 1:** COVID-19 Convalescent Plasma^a^ Donor Characteristics

| Characteristic | Count |
| --- | --- |
| Age |  |
| 18 – 29 | 2 |
| 30 – 39 | 1 |
| 40 – 49 | 3 |
| Gender |  |
| Male | 3 |
| Female | 3 |
| Race/Ethnicity |  |
| White | 6 |
| Blood Type |  |
| A+ | 2 |
| O+ | 4 |
| Days Since Diagnosis |  |
| 30 – 60 | 1 |
| 61 – 90 | 3 |
| 91 – 120 | 2 |

<sup>a</sup>One convalescent plasma unit was shipped frozen and the remaining 5 were shipped in liquid form on cold packs.

**Table 2:**
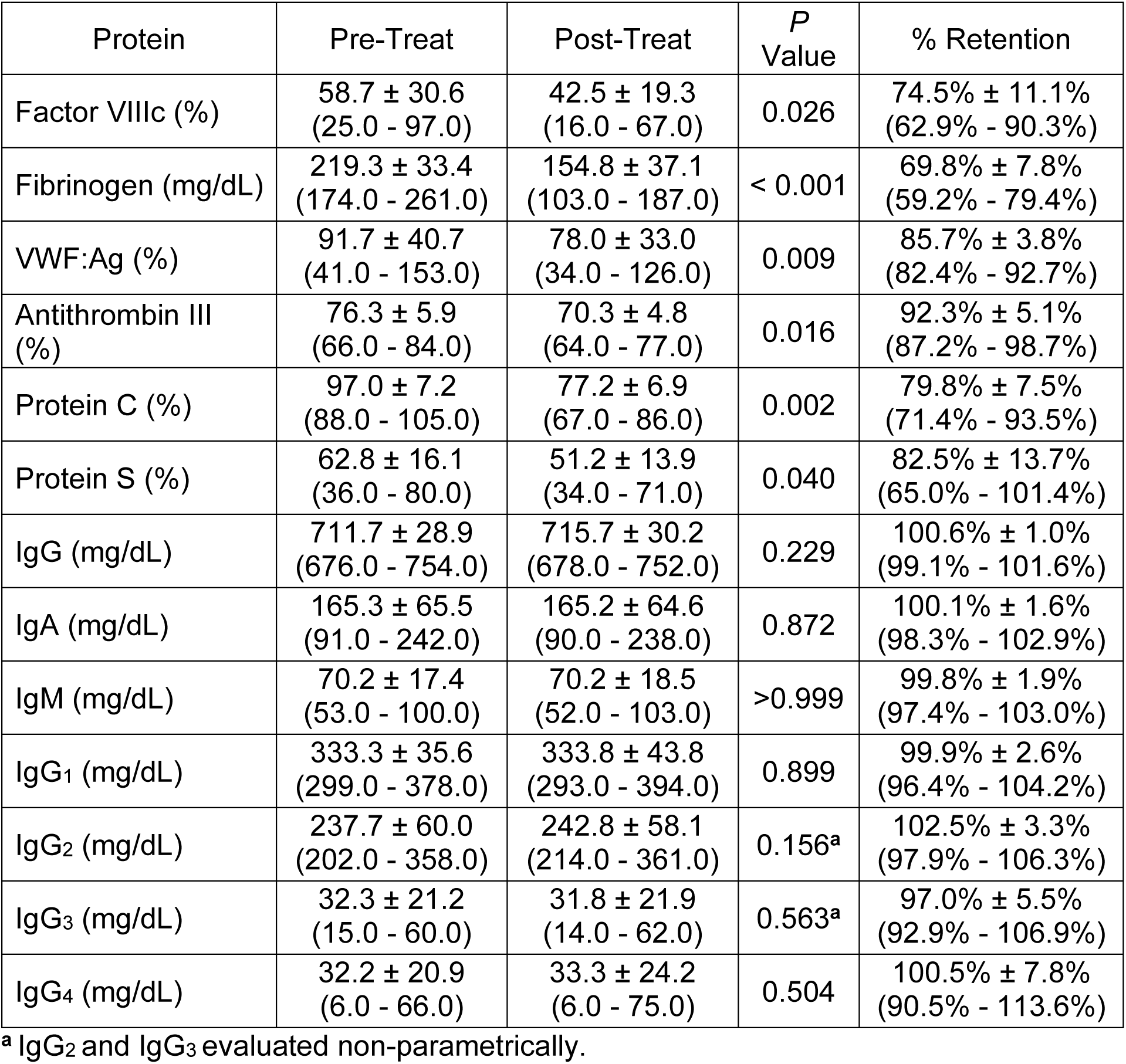
Protein Retention After R+UV PRT Treatment of COVID-19 Convalescent Plasma, mean ± 1 standard deviation (range)

| Protein | Pre-Treat | Post-Treat | *P*<br>Value | % Retention |
| --- | --- | --- | --- | --- |
| Factor VIIIc (%) | 58.7 ± 30.6<br>(25.0 - 97.0) | 42.5 ± 19.3<br>(16.0 - 67.0) | 0.026 | 74.5% ± 11.1%<br>(62.9% - 90.3%) |
| Fibrinogen (mg/dL) | 219.3 ± 33.4<br>(174.0 - 261.0) | 154.8 ± 37.1<br>(103.0 - 187.0) | < 0.001 | 69.8% ± 7.8%<br>(59.2% - 79.4%) |
| VWF:Ag (%) | 91.7 ± 40.7<br>(41.0 - 153.0) | 78.0 ± 33.0<br>(34.0 - 126.0) | 0.009 | 85.7% ± 3.8%<br>(82.4% - 92.7%) |
| Antithrombin III (%) | 76.3 ± 5.9<br>(66.0 - 84.0) | 70.3 ± 4.8<br>(64.0 - 77.0) | 0.016 | 92.3% ± 5.1%<br>(87.2% - 98.7%) |
| Protein C (%) | 97.0 ± 7.2<br>(88.0 - 105.0) | 77.2 ± 6.9<br>(67.0 - 86.0) | 0.002 | 79.8% ± 7.5%<br>(71.4% - 93.5%) |
| Protein S (%) | 62.8 ± 16.1<br>(36.0 - 80.0) | 51.2 ± 13.9<br>(34.0 - 71.0) | 0.040 | 82.5% ± 13.7%<br>(65.0% - 101.4%) |
| IgG (mg/dL) | 711.7 ± 28.9<br>(676.0 - 754.0) | 715.7 ± 30.2<br>(678.0 - 752.0) | 0.229 | 100.6% ± 1.0%<br>(99.1% - 101.6%) |
| IgA (mg/dL) | 165.3 ± 65.5<br>(91.0 - 242.0) | 165.2 ± 64.6<br>(90.0 - 238.0) | 0.872 | 100.1% ± 1.6%<br>(98.3% - 102.9%) |
| IgM (mg/dL) | 70.2 ± 17.4<br>(53.0 - 100.0) | 70.2 ± 18.5<br>(52.0 - 103.0) | >0.999 | 99.8% ± 1.9%<br>(97.4% - 103.0%) |
| IgG<sub>1</sub> (mg/dL) | 333.3 ± 35.6<br>(299.0 - 378.0) | 333.8 ± 43.8<br>(293.0 - 394.0) | 0.899 | 99.9% ± 2.6%<br>(96.4% - 104.2%) |
| IgG<sub>2</sub> (mg/dL) | 237.7 ± 60.0<br>(202.0 - 358.0) | 242.8 ± 58.1<br>(214.0 - 361.0) | 0.156<sup>a</sup> | 102.5% ± 3.3%<br>(97.9% - 106.3%) |
| IgG<sub>3</sub> (mg/dL) | 32.3 ± 21.2<br>(15.0 - 60.0) | 31.8 ± 21.9<br>(14.0 - 62.0) | 0.563<sup>a</sup> | 97.0% ± 5.5%<br>(92.9% - 106.9%) |
| IgG<sub>4</sub> (mg/dL) | 32.2 ± 20.9<br>(6.0 - 66.0) | 33.3 ± 24.2<br>(6.0 - 75.0) | 0.504 | 100.5% ± 7.8%<br>(90.5% - 113.6%) |

<sup>a</sup> IgG<sub>2</sub> and IgG<sub>3</sub> evaluated non-parametrically.

All 6 CCP units demonstrated binding to the SARS-CoV-2 RBD as well as the S1 and S2 subunits of the spike protein when assessed by ELISA using anti-IgG and anti-IgM secondary antibodies. The levels of IgM antibodies detected were generally lower and more variable than IgG antibodies, particularly for those targeted against the RBD, but normalized AUC values did not significantly differ between Pre-Treat and Post-Treat time points for either IgG or IgM at any of the binding sites (Figure 1). Similarly, SARS-CoV-2 neutralizing activity was detected by the PRNT assay in all of the Post-Collect and Pre-Treat CCP samples, though one unit was at the 1:20 threshold. The PRNT_50_ titer for one unit (370020801130) dropped by one dilution between Post-Collect and Pre-Treat, but all CCP units demonstrated stable PRNT_50_ titers when comparing Pre-Treat and Post-Treat samples (Table 3). Two units and one additional Pre-Treat sample tested negative by the RVPN assay, and estimated RVPN NT_50_ titers were variable (Table 4).

**Figure 1:**
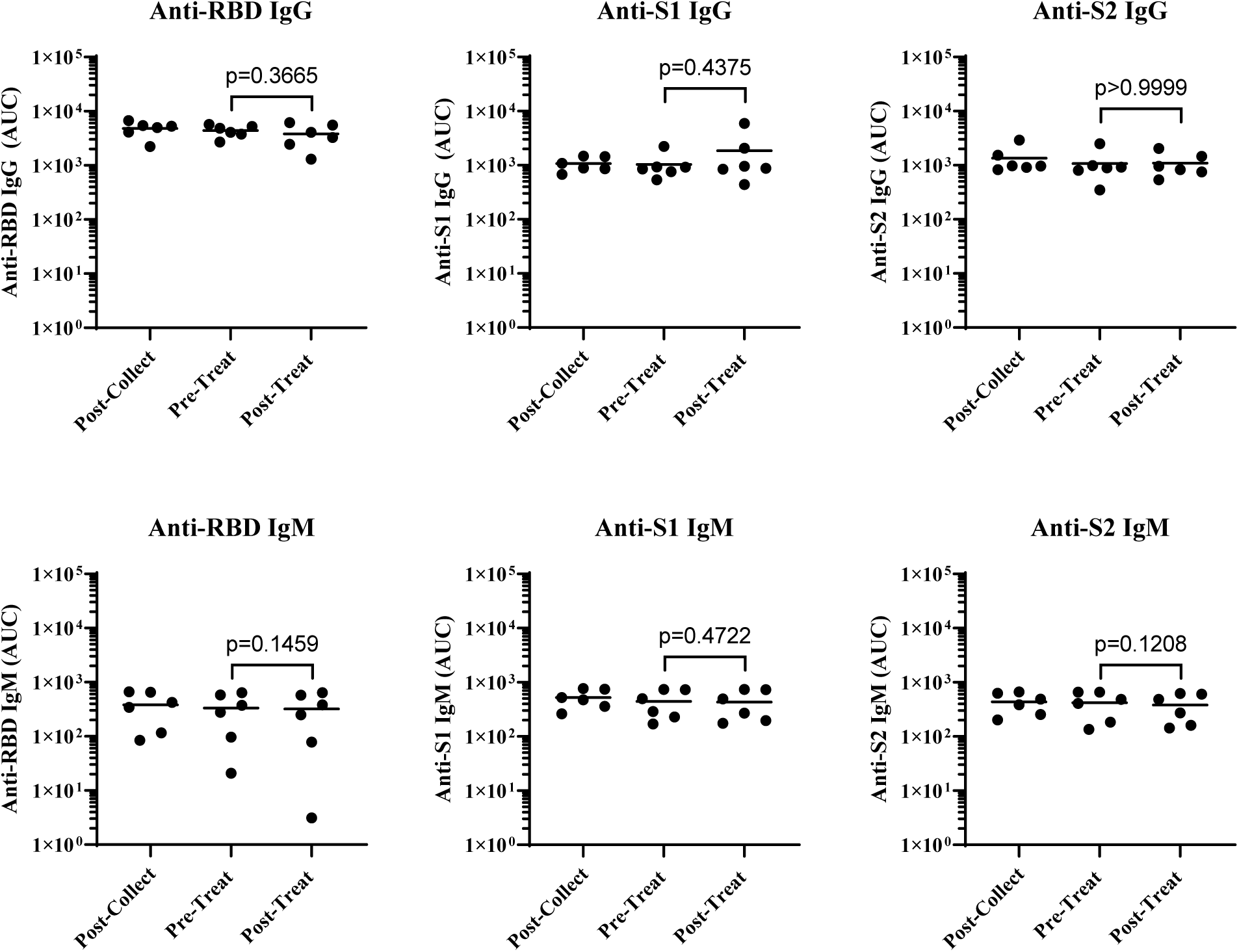
Plasma antibodies against SARS-CoV-2. ELISA results expressed as area under the curve (AUC) values based upon optical density at 450 nm (OD^450^) measurements over a range of plasma dilutions (Supplemental Figure 1).

**Table 3:** SARS-CoV-2 PRNT_50_ Limiting Dilution Titers

| CCP Unit ID | Post-Collect | Pre-Treat | Post-Treat |
| --- | --- | --- | --- |
| 370020800808 | 1:80 | 1:80 | 1:80 |
| 370020800898 | 1:40 | 1:40 | 1:40 |
| 370020800970 | 1:40 | 1:40 | 1:40 |
| 370020801002<sup>a</sup> | 1:20 | 1:20 | 1:20 |
| 370020801095 | 1:320 | 1:320 | 1:320 |
| 370020801130 | 1:80 | 1:40 | 1:40 |

<sup>a</sup>This unit is negative based upon a PRNT<sub>50</sub> threshold $\leq 1:20$ .

**Table 4:**
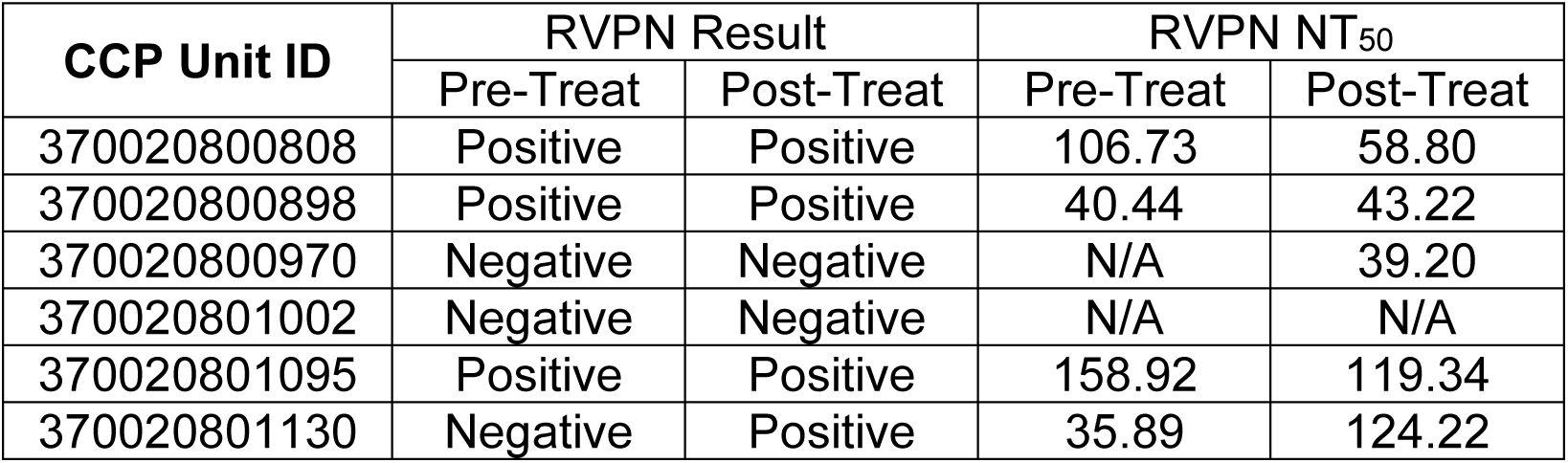
SARS-CoV-2 Pseudovirus Reporter Viral Particle Neutralization (RVPN) Assay

| CCP Unit ID | RVPN Result |  | RVPN NT<sub>50</sub> |  |
| --- | --- | --- | --- | --- |
|  | Pre-Treat | Post-Treat | Pre-Treat | Post-Treat |
| 370020800808 | Positive | Positive | 106.73 | 58.80 |
| 370020800898 | Positive | Positive | 40.44 | 43.22 |
| 370020800970 | Negative | Negative | N/A | 39.20 |
| 370020801002 | Negative | Negative | N/A | N/A |
| 370020801095 | Positive | Positive | 158.92 | 119.34 |
| 370020801130 | Negative | Positive | 35.89 | 124.22 |

## Discussion

This study evaluated the effect of R+UV PRT treatment on functional properties of CCP. A treatment effect upon coagulation factors was observed following R+UV treatment, but the reductions seen were consistent with previously published R+UV literature [18, 22-25]. Moreover, all PRT methods are known to degrade plasma proteins to varying degrees [26-29]. Minimal effects upon antibodies were demonstrated, from the very general immunoglobulin retention percentages to the more specific SARS-CoV-2 epitope binding measurements. Neutralizing antibody activity was similarly well-preserved, with the highest RVPN dilutions positive for neutralizing activity remaining the same after treatment and Pre-Treat and Post-Treat PRNT^50^ values being identical for all CCP units. The PRNT_50_ titer for one CCP unit dropped when comparing Post-Collect and Pre-Treat samples, which is likely an artifact of dilution with riboflavin solution during the R+UV PRT treatment process. The stability demonstrated in this study is consistent with previous assessments of antibody function in PRT-treated plasma [30, 31]. These data suggest that PRT treatment does not impair the passive immunity provided by CCP.

Some differences were observed between the results provided by the two assays for nAb activity. An additional unit was deemed negative for nAb activity and lower titers were reported for two units by the RVPN assay compared to the PRNT assay. The higher sensitivity of the PRNT assay may stem from greater susceptibility of the wild type virus to a more diverse set of antibodies or quaternary epitopes that cannot be replicated with the pseudovirus [32]. While the PRNT assay has higher sensitivity, working with live SARS-CoV-2 requires BSL-3 containment measures. The RVPN assay was developed to quantitatively measure SARS-CoV-2 neutralization titers safely in laboratory facilities typical of blood centers to select CCP units for therapeutic use [21]. RVPN NT_50_ values were quite variable and most likely were not representative of R+UV PRT treatment effects. Given the low titer of the CCP units evaluated in the study, the non-linear regression used to calculate the titer was based upon a limited non-zero dataset, thereby affecting the accuracy of the estimate. This would not be an issue at therapeutic antibody titers.

Importantly, the levels of IgG and IgM antibodies to specific viral proteins in the receptor binding domain (RBD) and spike proteins (S1 and S2) were maintained following treatment. These antibodies have been shown to have high virus neutralizing capacity. Robbiani et al. [19] demonstrated that despite variations in the levels of overall neutralizing antibodies in donors of convalescent plasma, the presence of these specific subsets of antibodies with potent antiviral activity correlated with improved clinical outcomes in patients receiving the convalescent plasma products. The data implies that maintenance of the level of these subsets of antibodies may correlate with clinical effectiveness more directly than measure of overall neutralizing antibody levels.

CCP is the most readily available source of anti-SARS-CoV-2 antibodies, and its use has been widely embraced as a treatment for COVID-19 while other antiviral therapies and vaccines are in development and can be widely deployed. The ability to safely utilize convalescent plasma in these settings, however, depends on the safety of the product collected from donors who may have experienced a period of immune compromise during acute phases of the disease. Exposure to a variety of transfusion-transmitted diseases during this period or reactivation of latent disease could introduce additional risk into the use of such products for therapeutic applications. PRT treatment of CCP may be seen as a prudent safety measure to mitigate the risk of possible co-infections known to be transmissible by transfusion. The ability to limit the risk of transfusion-transmitted co-infections is of particular importance in areas having a high prevalence of endemic disease, as is the case in many resource-limited settings. Local collection of CCP in these environments may be challenged by the need for apheresis infrastructure and cold chain requirements [33], though success in establishing a CP supply chain to support an EVD clinical trial in Guinea through the collaboration of international research consortia, government agencies, charitable foundations, and blood establishments has been described [34]. Since scale-up of such a system to serve the needs of the broader population for the COVID-19 pandemic is likely not feasible, whole blood (WB)-derived CCP or perhaps even convalescent WB may be more plausible where resources are limited. There is precedent for efficacious use of convalescent WB against EVD, and WB collection is far simpler to than plasmapheresis [35]. PRT systems to treat WB are available or in development, including the R+UV PRT system used to treat CCP in this study [36]. Although the preservation of antibody function in R+UV-treated WB was not evaluated in this study, R+UV treatment effects on plasma coagulation factors are similar to those reported herein [37].

Limitations of this study include the small sample size and the generally low anti-SARS-CoV-2 titers in the CCP units. In an Emergency Use Authorization for the use of CCP to treat hospitalized COVID-19 patients, the United States Food and Drug Administration defined high-titer CCP to be units with a signal-to-cutoff value of 12 or greater when tested by the Ortho VITROS SARS-CoV-2 IgG test, which corresponds to an ID_50_ titer cutoff of 250 using a SARS-CoV-2 neutralization assay similar to the PRNT [38]. The six CCP units evaluated in this study were collected specifically for research at a time when blood centers were urgently calling for therapeutic CCP donations. It is possible that the donors providing research CCP units were unable to donate therapeutic units due to low antibody titers or other donor deferral factors. Despite the low titers, the various antibody assays performed in this study consistently demonstrated stability between pre- and post-treatment samples, whether testing for retention, epitope binding, or neutralizing activity.

## Conclusions

With the worldwide need for treatment options to address the COVID-19 pandemic, CCP is an expedient therapeutic option that can be implemented globally, whether in resource-rich or resource-limited environments. The addition of PRT may be warranted to address possible co-infections in regions experiencing a high prevalence of endemic transfusion-transmissible diseases, but conservation of the passive immunity conveyed through CCP must be ensured. Based upon this small study there is no indication that R+UV PRT treatment compromises SARS-CoV-2 nAb function in COVID-19 convalescent plasma.

## Data Availability

N/A

## Acknowledgements

The authors wish to acknowledge Key Biologics for contributing COVID-19 convalescent plasma for this work. We also thank BEI for providing the SARS-CoV-2 utilized in these studies. The reagent was deposited by the Centers for Disease Control and Prevention and obtained through BEI Resources, NIAID, NIH: SARS-Related Coronavirus 2, Isolate USA-WA1/2020, NR-52281. Larry Dumont at Vitalant Research Institute kindly provided critical review. Lastly, we acknowledge the contributions of Aja Anderson for coordinating sample shipments among laboratories and Kalen Quintanar for performing coagulation factor assays.

## Disclaimer

The views expressed in this manuscript are the authors’ own views and do not represent an official position of their institutions or the organizations funding this research.

---

**Supplemental Figure 1:**
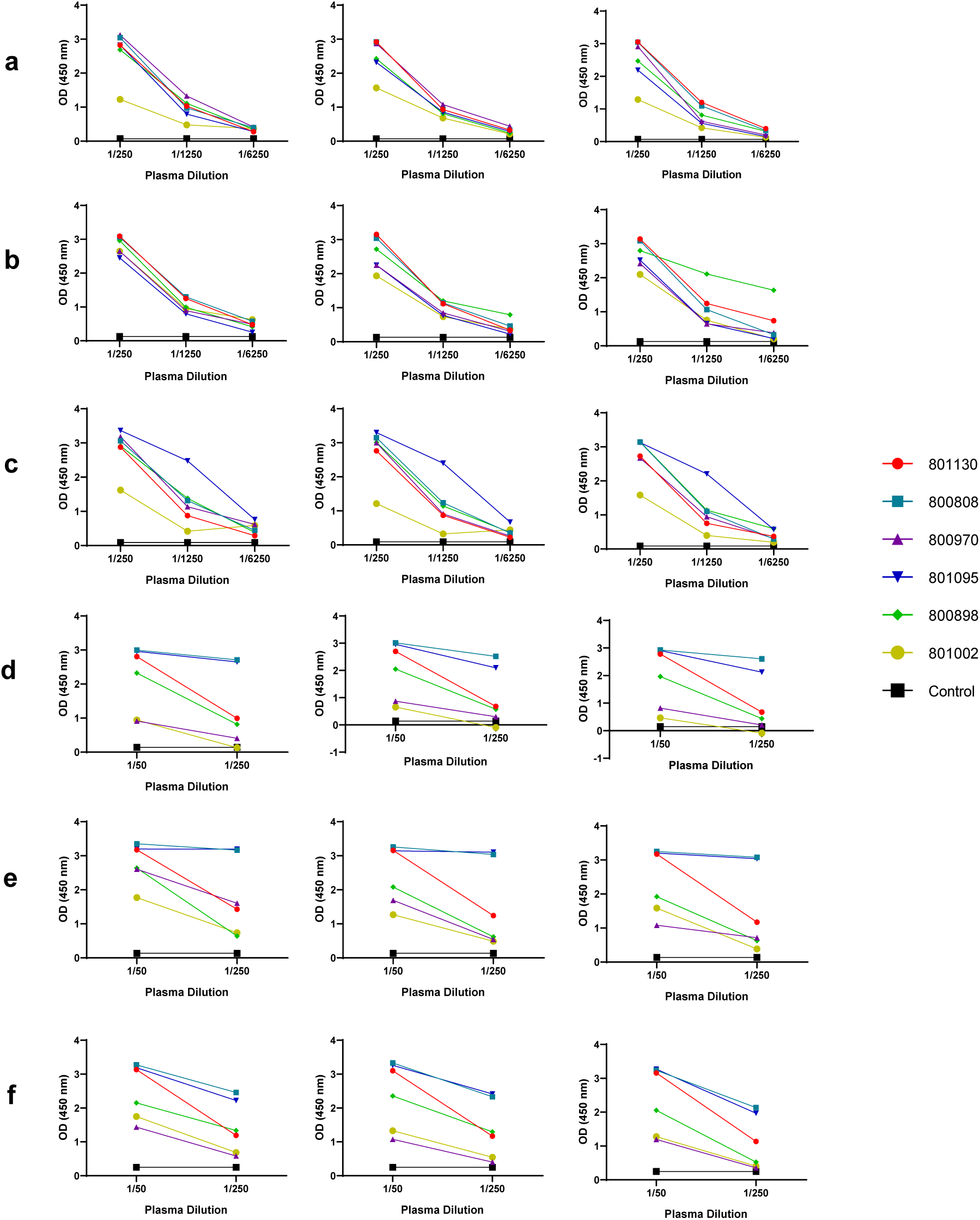
Optical density at 450 nm (OD450nm). Plasma dilutions used to calculate area under the curve (AUC) for the 6 convalescent plasma units and a normal control at Post-Collect (left), Pre-Treat (middle) and Post-Treat (right) time points; (a) anti-RBD IgG; (b) anti-S1 IgG; (c) anti-S2 IgG; (d) anti-RBD IgM; (e) anti-S1 IgM; (f) anti-S2 IgM

## Notes

### Competing Interest Statement

S.Y. and S.M. are employees of Terumo Blood and Cell Technologies, the manufacturer of the pathogen reduction technology described in this article. L.H., T.D., M.H.D. and R.G. have no conflicts of interest to declare.

### Clinical Trial

This study was not a clinical trial; it was an in vitro study of human COVID-19 convalescent plasma

### Funding Statement

This study was funded by Terumo Blood and Cell Technologies

### Author Declarations

COVID-19 convalescent plasma was collected at Key Biologics under protocol "COLLECTION OF BLOOD COMPONENTS BY LEUKAPHERESIS FROM PATIENTS POST SARS-COV-2 INFECTION FOR SUPPORT OF RESEARCH," approved by Advarra IRB. Confirmation of ethics approval for CCP collection was provided to our research collaborators by Edward P. Scott, MD, Principal Investigator at Key Biologics. CCP unit ID numbers are not traceable to donor identity outside of Key Biologics. Ethical approval for the study was provided by the Colorado State University Research Integrity & Compliance Review Office, Fort Collins, CO, under the "Indications against Highly Pathogenic Agents for a Transportable Pathogen Reduction and Blood Safety System for Whole Blood, Platelets and Plasma" protocol. Per the IRB review it was determined that the activity did not meet the requirements of the federal definition of human subject research.

